# COA5 has an essential role in the early stage of mitochondrial complex IV assembly

**DOI:** 10.1101/2024.08.27.24312374

**Authors:** Jia Xin Tang, Alfredo Cabrera-Orefice, Jana Meisterknecht, Lucie S. Taylor, Geoffray Monteuuis, Maria Ekman Stensland, Adam Szczepanek, Karen Stals, James Davison, Langping He, Sila Hopton, Tuula A. Nyman, Christopher B. Jackson, Angela Pyle, Monika Winter, Ilka Wittig, Robert W. Taylor

**Author notes:** Department of NanoBiophotonics, Max Planck Institute for Multidisciplinary Sciences, Göttingen, Germany (Jia Xin Tang).

## Abstract

Pathogenic variants in cytochrome *c* oxidase assembly factor 5 (COA5), a proposed complex IV (CIV) assembly factor, have been shown to cause clinical mitochondrial disease with two siblings affected by neonatal hypertrophic cardiomyopathy manifesting a rare, homozygous *COA5* missense variant (NM_001008215.3: c.157G>C, p.Ala53Pro). The most striking observation in the affected individuals was an isolated impairment in the early stage of mitochondrial CIV assembly. In this study, we report an unrelated family in who we have identified the same *COA5* variant with patient-derived fibroblasts and skeletal muscle biopsies replicating an isolated CIV deficiency. A CRISPR/Cas9-edited homozygous *COA5* knockout U2OS cell line with similar biochemical profile was generated to interrogate the functional role of the human COA5 protein. Mitochondrial complexome profiling pinpointed a role for COA5 in early CIV assembly, more specifically, its involvement in the stage between MTCO1 maturation and the incorporation of MTCO2. We therefore propose that the COA5 protein plays an essential role for the biogenesis of MTCO2 and its integration into the early CIV assembly intermediate for downstream assembly of the functional holocomplex.

## Introduction

Mitochondria synthesise cellular energy in the form of adenosine triphosphate (ATP) via oxidative phosphorylation (OXPHOS), comprising four respiratory chain complexes and the F_1_F_O_ ATP synthase. Cytochrome *c* oxidase (COX), also known as complex IV (CIV), is the terminal electron acceptor of the respiratory chain which catalyses the reduction of molecular oxygen to water. CIV couples this redox reaction to the translocation of protons across the inner mitochondrial membrane, thus contributing to the generation of the proton-motive force harnessed by the F_1_F_O_ ATP synthase to generate ATP.

CIV is comprised of 14 protein subunits of dual genetic origin; the three core subunits (MTCO1, MTCO2 and MTCO3) are all mitochondrially-encoded while the remaining subunits are encoded by the nuclear genome (Kadenbach, 2017; Wikström *et al*, 2018). Remarkably, CIV possesses two redox active copper centres (binuclear Cu_A_ and mononuclear Cu_B_ centres) and two haem groups (haem *a* and haem *a_3_*) (Wikström *et al*., 2018). These redox active cofactors are crucial for electron transfer within CIV which entails: (i) the receipt of electrons from reduced cytochrome *c* by the Cu_A_ centre in the MTCO2 subunit, (ii) subsequent delivery of the electron by the haem *a* group in membrane-spanning MTCO1 subunit to (iii) the oxygen-reducing haem *a_3_*-Cu_B_ centre (Belevich *et al*, 2006; Kirchberg *et al*, 2012; Muramoto *et al*, 2010). These reactions result in the pumping of a total of four protons per oxygen molecule into the mitochondrial intermembrane space (IMS).

As a consequence, an intricate assembly machinery has been generally described for complex IV on a modular basis centred around the three catalytic subunits: MTCO1, MTCO2 and MTCO3 with over 20 unique assembly factors of CIV having been characterised to date (Signes & Fernandez-Vizarra, 2018; Vidoni *et al*, 2017; Watson & McStay, 2020). These assembly factors are not only involved in the sequential incorporation of the protein subunits but also crucial for auxiliary processes such as translational regulation, protein stabilisation as well as the insertion of cofactors and prosthetic groups (Povea-Cabello *et al*, 2024; Watson & McStay, 2020).

The cytochrome *c* oxidase assembly factor 5 (*COA5*) gene (RefSeq: NM_001008215.3), previously denoted as *C2orf64*, was first reported in humans when a homozygous missense variant (c.157G>C, p.Ala53Pro) in this gene was shown to cause mitochondrial disease. Biochemical studies of patient-derived fibroblasts revealed isolated COX deficiency, more specifically the accumulation of CIV assembly intermediates and decreased levels of fully assembled CIV holocomplexes, leading the authors to hypothesise that COA5 is involved in the early stages of CIV assembly (Huigsloot *et al*, 2011). However, earlier studies carried out using the yeast orthologue of human COA5 protein, Pet191, also suggested a putative role in CIV assembly but with no impact on COX translation and copper metalation of the protein (Khalimonchuk *et al*, 2008; McEwen *et al*, 1993; Tay *et al*, 2004). Interestingly, contradicting evidence have been published with regards to the mitochondrial localisation of the Pet191 protein despite being a member of the twin CX_9_C protein family that are often found to be dependent on the Mitochondrial Intermembrane space Import and Assembly (MIA) pathway (Bragoszewski *et al*, 2013; Khalimonchuk *et al*., 2008).

Here we present an unrelated family in which a clinically affected child harbours the identical *COA5* missense variant, identified by trio whole exome sequencing (Longen *et al*). We generated a CRISPR/Cas9-mediated *COA5* knockout (*COA5*^KO^) cell line to elucidate the role of COA5 in CIV assembly and its implications on mitochondrial health and disease, utilising an array of biochemical tools including mitochondrial complexome profiling.

## Results

### Clinical summary and genomic studies

A female neonate born to second cousin parents presented with tachypnoeic episodes on the first day of life. Blood lactate was elevated at variable levels between 2.5 to 11 mmol/L. Clinical examination detected hyperdynamic precordium with loud second heart sound and subsequent echocardiography indicated significant biventricular hypertrophic cardiomyopathy with septal hypertrophy and non-compaction appearance of the myocardium. Due to worsening respiratory distress, she was intubated and given inotropic support after birth before being discharged. Liver function tests were abnormal with increased echogenicity of liver on ultrasound. Neuroimaging of the brain was normal. Focused metabolic biochemical investigations identified elevated urinary lactate and ethylmalonic acid. Clinical care was directed to symptomatic management only when a presumptive diagnosis of mitochondrial disorder with hypertrophic cardiomyopathy was made having identified evidence of significant complex IV activity deficiency on muscle biopsy as detailed below. She was readmitted several weeks later with further deterioration of respiratory distress concurrent with a rhinovirus infection, poor cardiac function and severe lactic acidosis and sadly passed away.

Molecular genetic testing eliminated pathogenic mitochondrial DNA (mtDNA) variants following a complete analysis of the mitochondrial genome. Trio whole exome sequencing conducted at the Exeter Genomics Laboratory identified a previously reported *COA5* variant (c.157G>C, p.Ala53Pro), confirming both parents to be heterozygous carriers (**Fig 1A**) (Chen *et al*, 2023; Huigsloot *et al*., 2011). The homozygous *COA5* missense variant has been recorded on ClinVar as a pathogenic variant associated with isolated COX deficiency (https://www.ncbi.nlm.nih.gov/clinvar/variation/31087/). The c.157G>C *COA5* variant causes an amino acid change from alanine to proline at position 53 of the COA5 protein, which is located within the CX_9_C domain that has suggested linkage to mitochondrial protein localisation (Gladyck *et al*, 2021) (**Fig 1B**). The alanine residue is not conserved across species and is only shared between human and zebrafish (**Fig 1B**). Whilst the application of several *in silico* pathogenicity prediction tools suggested the c.157G>C *COA5* variant to be damaging, the REVEL meta-predictor score was below the recommended threshold for this variant, necessitating further study (Ioannidis *et al*, 2016).

**Figure 1.**
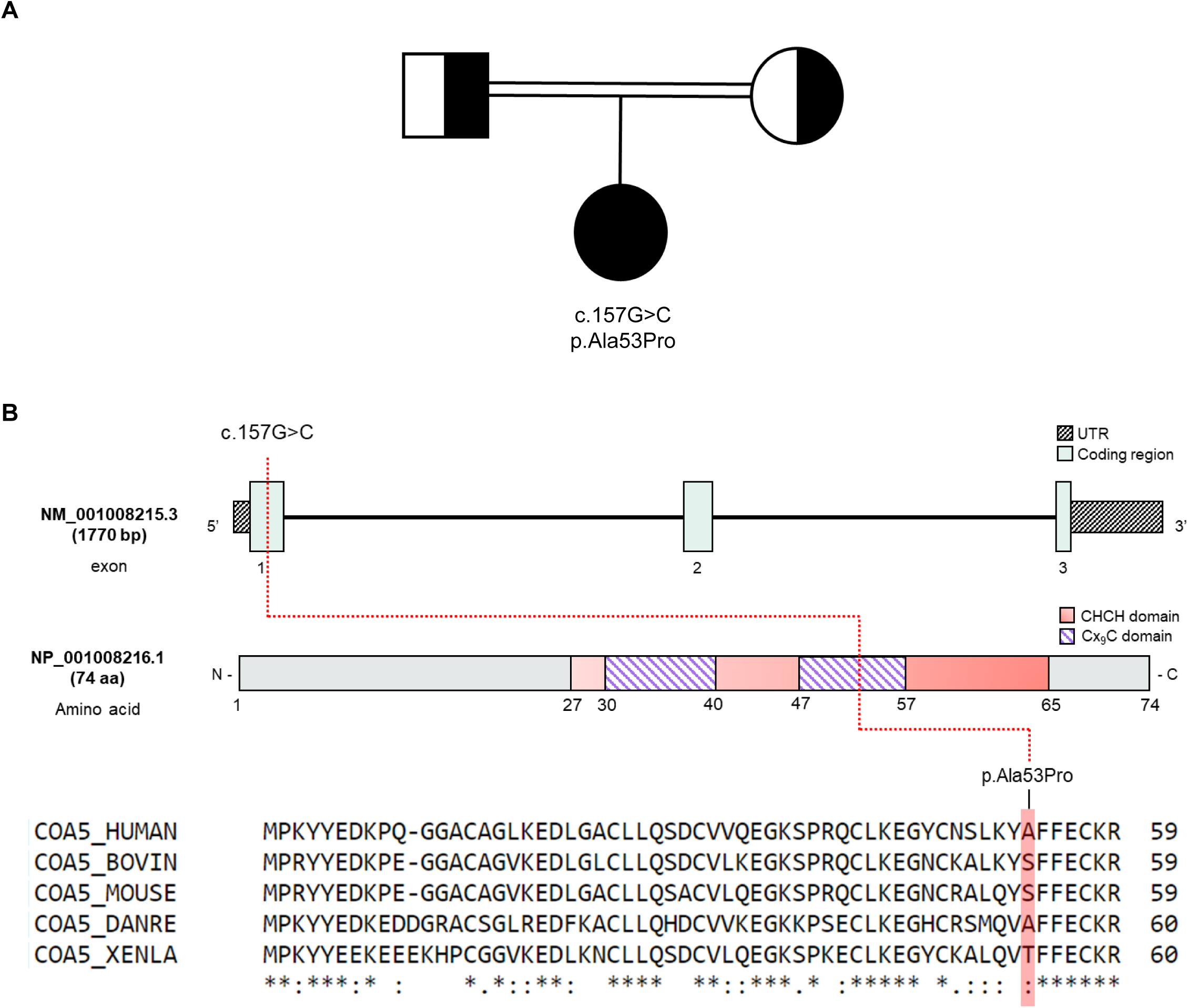
Molecular genetics of the *COA5* variant. **(A)** Partial family pedigree of *COA5* patient showing segregation of the *COA5* variant. **(B)** Schematic representation of the COA5 gene (Ensembl) and its encoded protein (InterPro and UniProt) illustrating the nucleotide or amino acid impacted by the *COA5* variant. The coiled coil-helix-coiled coil helix (CHCH) domain containing twin CX_9_C motifs (purple stripes) were illustrated in salmon pink. Amino acid residues at the position affected by the *COA5* variant across different species were highlighted.

### Patient-derived muscle biopsies and fibroblasts displayed isolated complex IV deficiency

Histochemical analysis of skeletal muscle cryosections from the patient indicated a moderate CIV deficiency in all fibres (**Fig 2A**). This was corroborated by quadruple OXPHOS immunofluorescence assay which showed a marked loss of MTCO1 immunoreactivity (**Fig 2B**) while NDUFB8 (complex I subunit) protein levels were normal (Ahmed *et al*, 2017). The direct measurement of respiratory chain enzyme activities in patient-derived fibroblasts also indicated a severe and isolated complex IV deficiency in the *COA5* patient muscle sample (**Fig 2C**).

**Figure 2.**
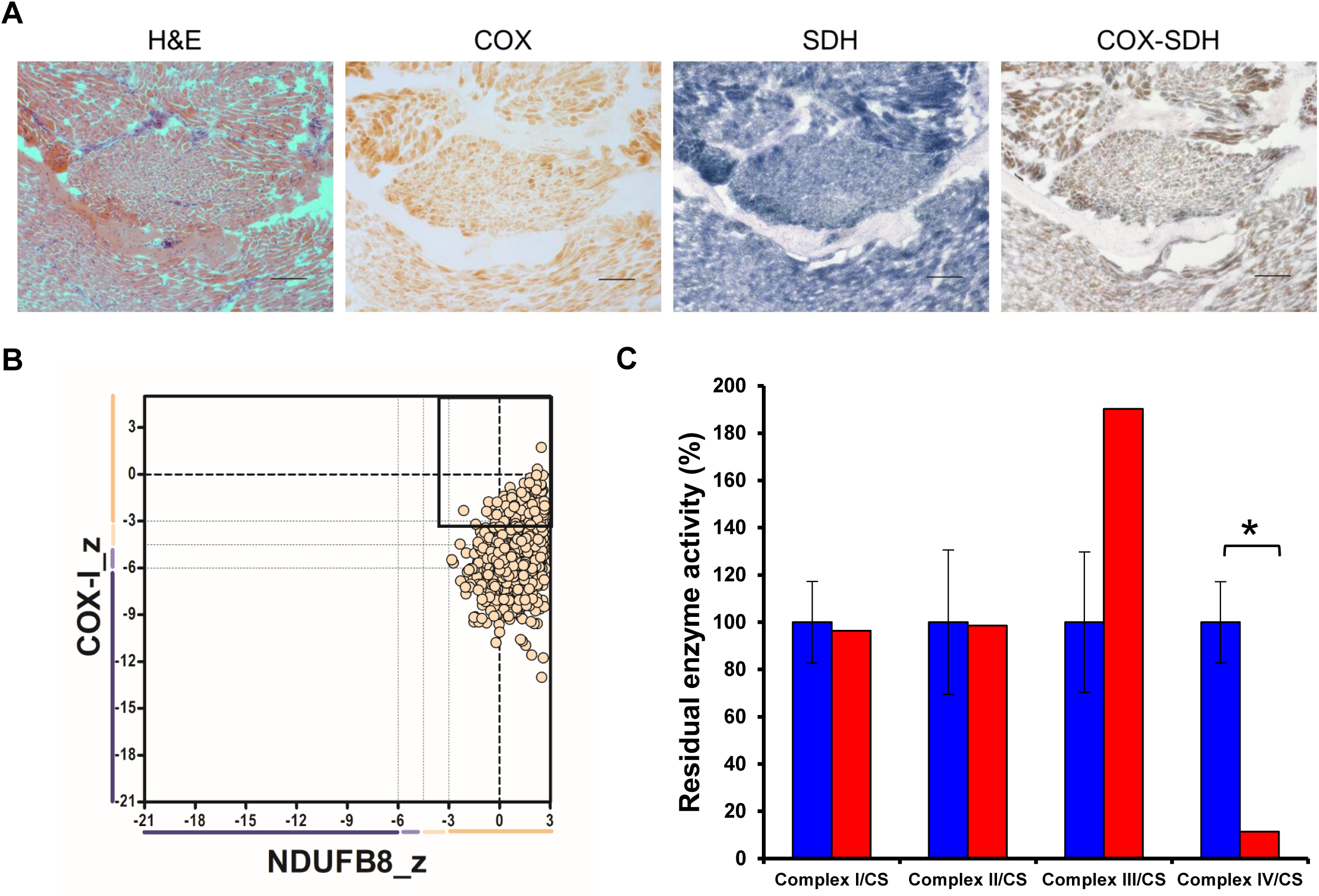
Histopathology findings of patient with *COA*5 variant. **(A)** Histochemical analysis of skeletal muscle section from patient with haematoxylin and eosin (H&E), cytochrome *c* oxidase (COX), succinate dehydrogenase (SDH) and sequential COX-SDH histochemistry. Scale bar = 100 µm; magnification at 10x**. (B)** Quadruple OXPHOS immunofluorescent assay of single skeletal muscle fibres from patient showing immunoreactivity against complex I subunit, NDUFB8 (x-axis) and complex IV subunit, MTCO1 (y-axis) normalised with porin expression as mitochondrial mass marker. Each dot corresponds to a single muscle fibre and beige colour corresponds to normal mitochondrial mass. Muscle fibres with a z-score of less than −3 standard deviation are considered deficient. Bold dashed lines represent the mean expression level in healthy muscle fibres. **(C)** Spectrophotometric measurement of OXPHOS enzyme activities in patient fibroblasts (in red) compared to mean activities in age-matched controls (in blue) shown as 100%. All measurements were normalised to citrate synthase (CS) activity. Error bars represent standard deviations of enzyme activities in control fibroblasts *(n=8)*. Respiratory chain enzyme activities in the patient which exceed control range are marked with an asterisk (*).

### Implications of the homozygous COA5 variant on the steady-state level and assembly of complex IV

To characterise the pathogenicity of the *COA5* missense variant, SDS-PAGE and BN-PAGE were used to delineate its impact on OXPHOS protein steady-state levels and assembly. Decreased steady-state levels were only observed in CIV subunit, MTCO2 while other OXPHOS complex subunits were unaffected in whole cell lysates of both patient-derived fibroblasts and skeletal muscle biopsy (**Figs 3A** and **3C**). Likewise for BN-PAGE, only complex IV assembly was impaired in patient-derived fibroblasts and skeletal muscle whereas the remaining four OXPHOS complexes (CI, CII, CIII and CV) were unaffected, similar to age-matched controls (**Figs 3B** and **3D**). These observations were further supported by proteomic analysis of the immortalised patient fibroblasts which highlighted the isolated CIV deficiency. CIV protein subunits exhibited a statistically significant decrease in protein abundance up to almost four-fold while CI, CII, CIII and CV protein abundance remained unchanged (**Fig 3E**).

**Figure 3.**
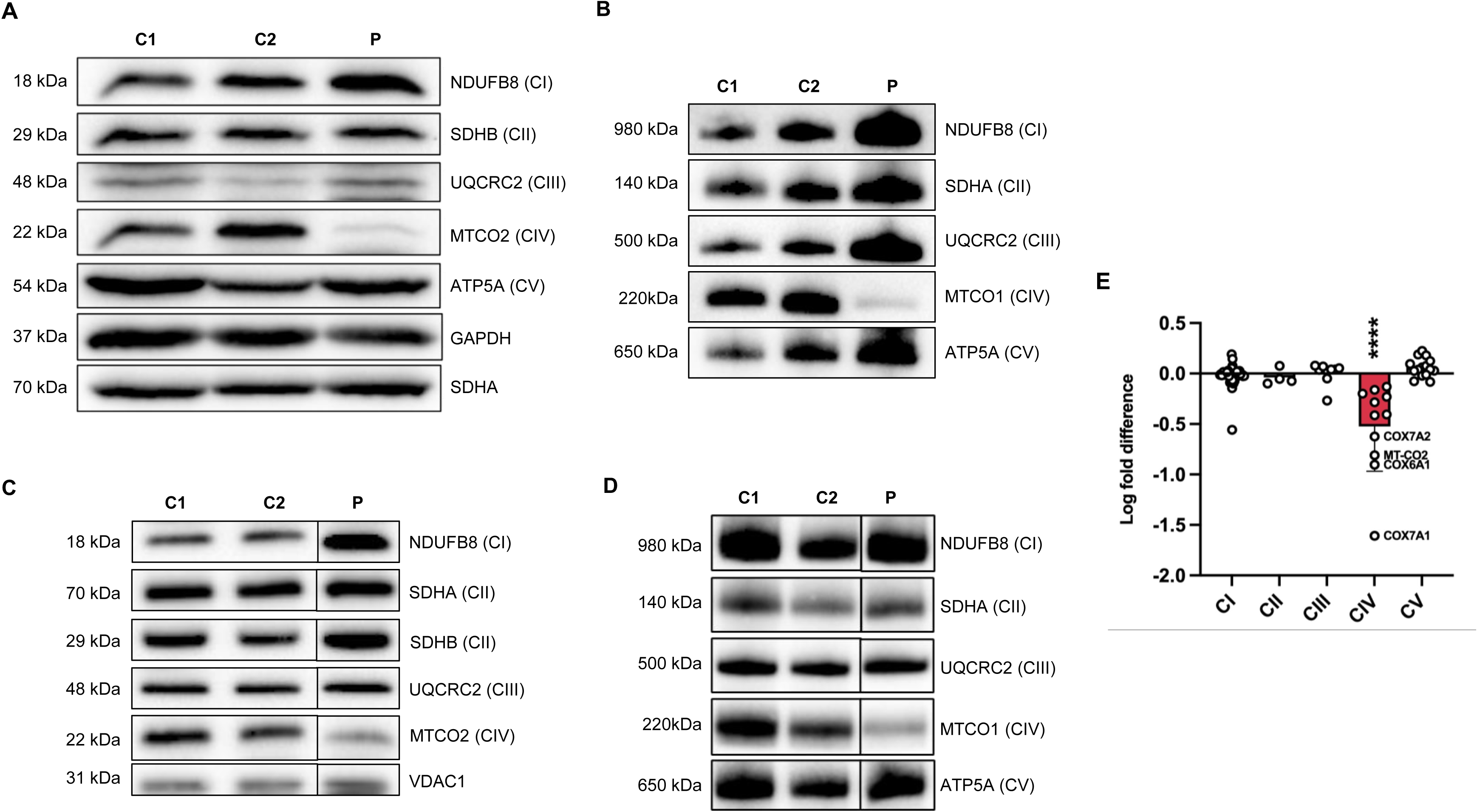
Immunoblotting analyses of COA5 patient-derived fibroblasts and skeletal muscle biopsies. **(A)** SDS-PAGE and immunoblotting analysis of whole protein lysates from *COA5* patient fibroblasts (P) and age-matched controls (C1 and C2) showing steady-state levels of OXPHOS complex subunits. GAPDH and SDHA were used as loading controls *(n=3)*. **(B)** BN-PAGE analysis of mitochondrial-enriched proteins from patient (P) against age-matched controls (C1 and C2) immunodetected against specific OXPHOS complex subunits. SDHA was used as loading control *(n=3)*. **(C)** Western blot analysis of protein extracts from skeletal muscle sections of patient (P) and controls (C1 and C2) with VDAC1 as loading control *(n=1)*. **(D)** BN-PAGE analysis of skeletal muscle mitochondrial extracts derived from patient (P) and age-matched controls (C1 and C2) detecting all five OXPHOS complex subunits with SDHA as loading control *(n=1)*. **(E)** Label-free proteomic profiling of immortalised patient fibroblasts with log-fold change in abundance compared to control. Each data point corresponds to the mean of quadruplicate measurements of identified OXPHOS complex subunits (40 out of 44 CI subunits, 3 out of 4 CII subunits, 9 out of 10 CIII subunits, 11 out of 19 CIV subunits and 15 out of 20 CV subunits were analysed.)

### Functional characterisation of COA5 in a CRISPR/Cas9 knockout cell line

Both the primary and immortalised patient fibroblasts harbouring the homozygous c.157G>C *COA5* variant displayed arrested cell growth in culture, likely owing to the severe CIV deficiency and therefore making further experimentation challenging in primary fibroblasts. To enable in-depth characterisation of the functional role and the implicated pathogenicity of *COA5*, CRISPR/Cas9 gene editing was utilised to generate a homozygous *COA5* knockout (*COA5*^KO^) in the immortalised human U2OS cell line for a more stable cellular model system. The *COA5*^KO^ cell line generated contained a homozygous 7 base pair deletion in the *COA5* gene (c.287_290+3del, p.Val61del) as verified by Sanger sequencing (**Figure EV1)**. Western blot analyses of the *COA5*^KO^ cell line also confirmed an isolated CIV defect in terms of protein steady-state level (**Fig 4A-B**) as well as OXPHOS complex assembly (**Fig 4C**), successfully mimicking the biochemical phenotype observed in patient-derived biopsies. Interestingly, an accumulation of the complex II subunit, SDHA protein at around 70 kDa was also observed on BN-PAGE analysis of the *COA5*^KO^ cell line which was absent in the isogenic control (**Fig 4C**).

**Figure 4.**
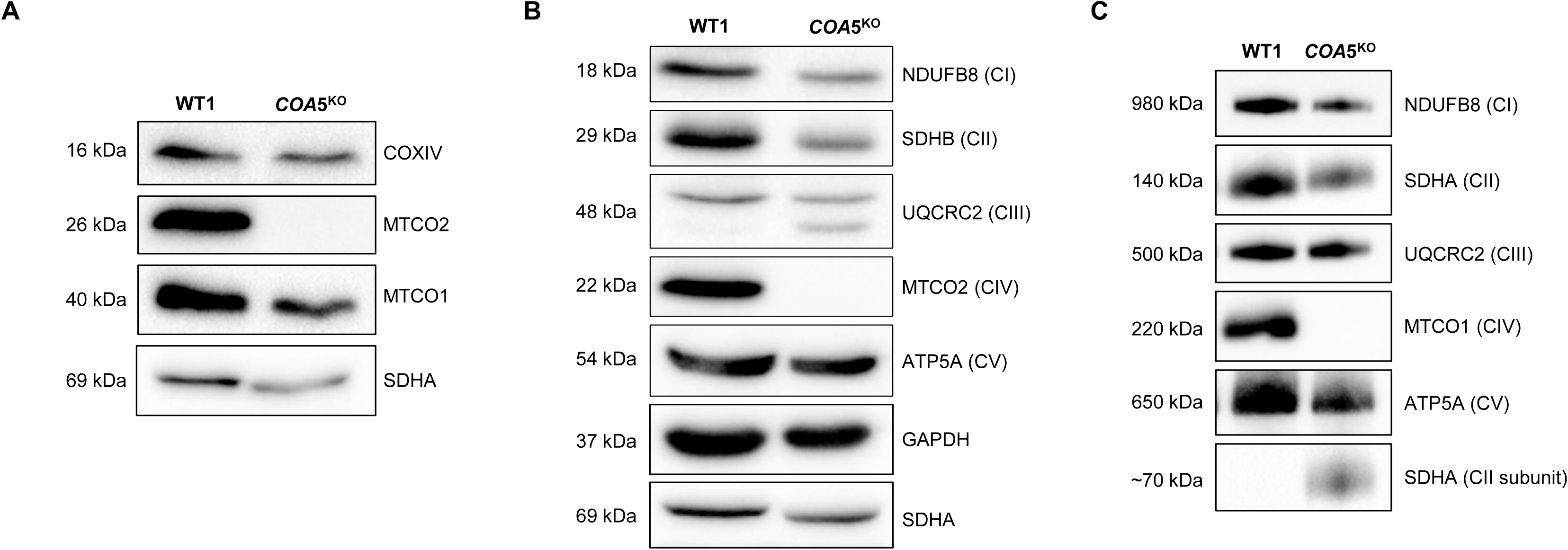
Western Blot analyses of COA5*^KO^* cell line against isogenic control. SDS-PAGE of whole cell lysates against **(A)** complex IV subunits including MTCO1, MTCO2 and COXIV *(n=2)* and **(B)** protein subunits of all five OXPHOS complexes using OXPHOS cocktail antibody *(n=2)*. GAPDH and/or SDHA were used as loading controls. **(C)** BN-PAGE of mitochondrial-enriched lysates immunoblotted against all five OXPHOS complexes with SDHA as loading control *(n=3)*.

### Mitochondrial Complexome Profiling of COA5^KO^ cell line

To define the functional involvement of COA5 in CIV assembly, mitochondrial complexome profiling, which combines BN-PAGE and tandem mass spectrometry (LC-MS/MS), was used to determine the presence and arrangement of the OXPHOS system and related protein complexes (Alahmad *et al*, 2020; Alston *et al*, 2018; Cabrera-Orefice *et al*, 2022; Lobo-Jarne *et al*, 2020). This technique enables visualisation of the specific stage of CIV assembly impacted when COA5 is absent using the *COA5*^KO^ cell line and potentially uncovering interacting partners of the COA5 protein to elucidate its functional role.

First, complexome profiling (**Fig 5**) confirmed our result on BN-PAGE_(**Fig 4**). An increased abundance of respiratory supercomplexes containing complexes I and III_2_ (S_0_: I+III_2_) was observed in the *COA5*^KO^ cell line (**Fig 5, right panel**). As observed in BN-PAGE analysis of the *COA5*^KO^ mitochondrial extracts (**Fig 4C**), accumulation of the SDHA subunit of CII was also detected by complexome profiling at a molecular size of approx 80 −100 kDa (native calibration of soluble complexes), but not in the isogenic control (highlighted in orange box in **Fig EV2, right panel**). Interesting, SDHAF1 and SDHAF2 comigrate at the same range indicating an assembly intermediate (**Fig EV3, right panel, orange box)**. When assessing complex III subunits, a complete loss of the supercomplexes III_2_+IV (S_0_) was observed in the *COA5*^KO^ cell line (**Fig 5**). Of all the OXPHOS complexes, only complex V content and assembly were unaffected (**Fig 5**).

**Figure 5.**
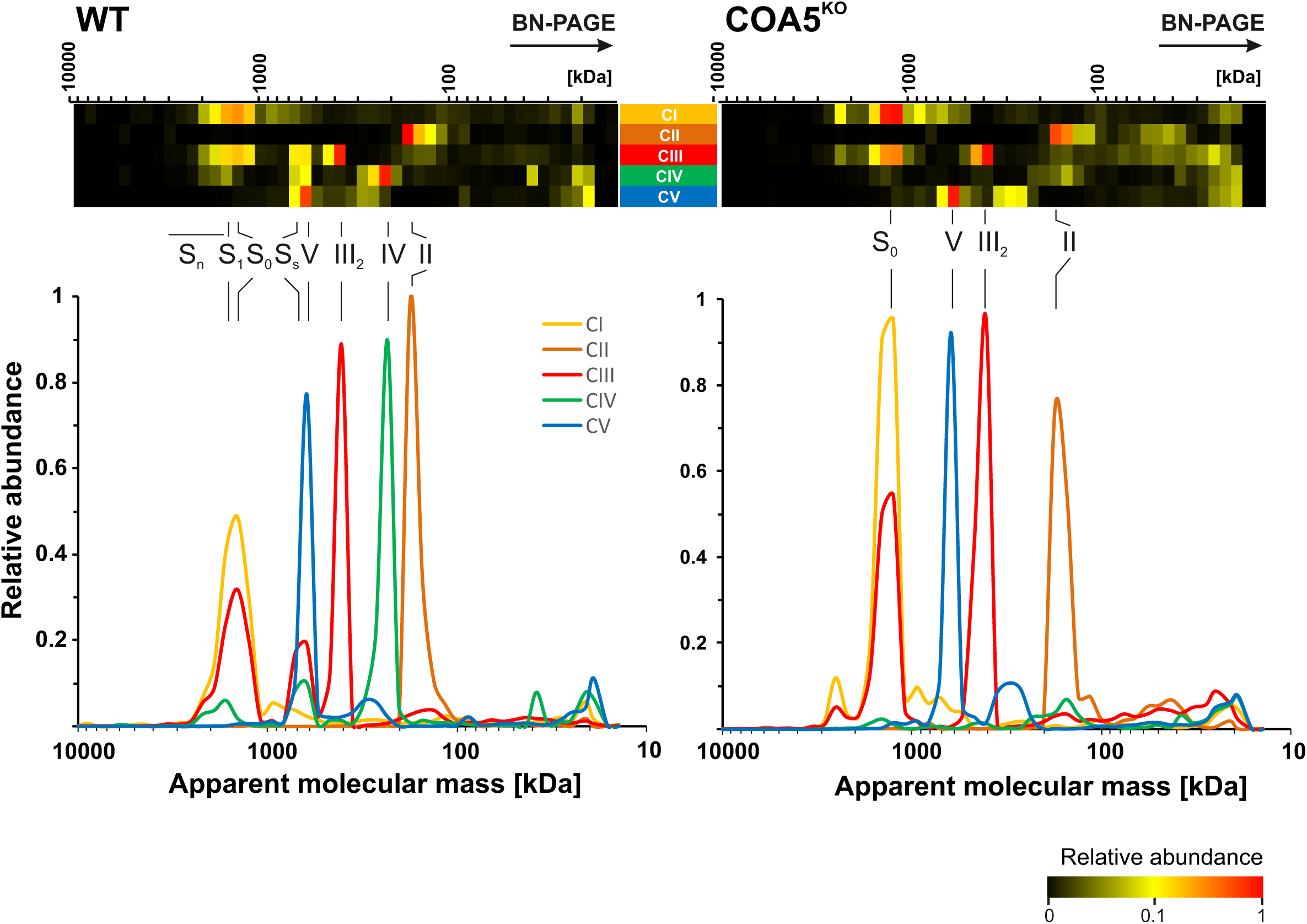
Complexome profiling identified decreased complex IV levels in COA5^KO^ compared to wildtype (COA5^WT^) isogenic control cell line. Isolated mitochondrial membranes were solubilised with digitonin and protein complexes were separated by BN-PAGE followed by quantitative mass spectrometric analysis. The quantitative data of identified individual subunits were summed up for each OXPHOS complex and normalised to maximum appearance between both cell lines. The data are presented as heat maps and 2D-plots, corresponding to protein components of individual OXPHOS complexes. The size of the complexes ranges from 10000 kDa to 10 kDa (from left to right). The corresponding complexes were highlighted above the heatmap and 2D-plots: fully assembled complex IV holocomplexes (IV), complex III dimer (III_2_), supercomplexes of complex III dimers and complex IV (S_S_) and supercomplexes containing complex I, complex III dimer (S_0_) and complex IV (S_1_).

Next, we had a closer look to complex IV subunits and assembly factors. We noted a complete loss of the COA5 protein as the respective 12 kDa protein was not detected in the knockout cell line but clearly present in the isogenic control as highlighted (**Fig 6A, highlighted in yellow**). More importantly, the accumulation of early CIV assembly intermediates (**Fig 6A, right panel, green boxed subunits and assembly factors**) and the loss of fully assembled CIV holocomplexes were observed in contrast to the isogenic control cell line (**Fig 6A, left panel**). Interestingly, subunits of the MTCO1 and MTCO2 modules accumulate in this intermediate, suggesting that despite the lack of subunits in the individual modules, further assembly is already taking place. This provides supporting evidence to the putative role of COA5 as an assembly factor in early stages of complex IV biogenesis and the isolated CIV defect resulting from the loss of COA5 protein as observed in biochemical studies of patient-derived cells and tissues (**Fig 3**).

**Figure 6.**
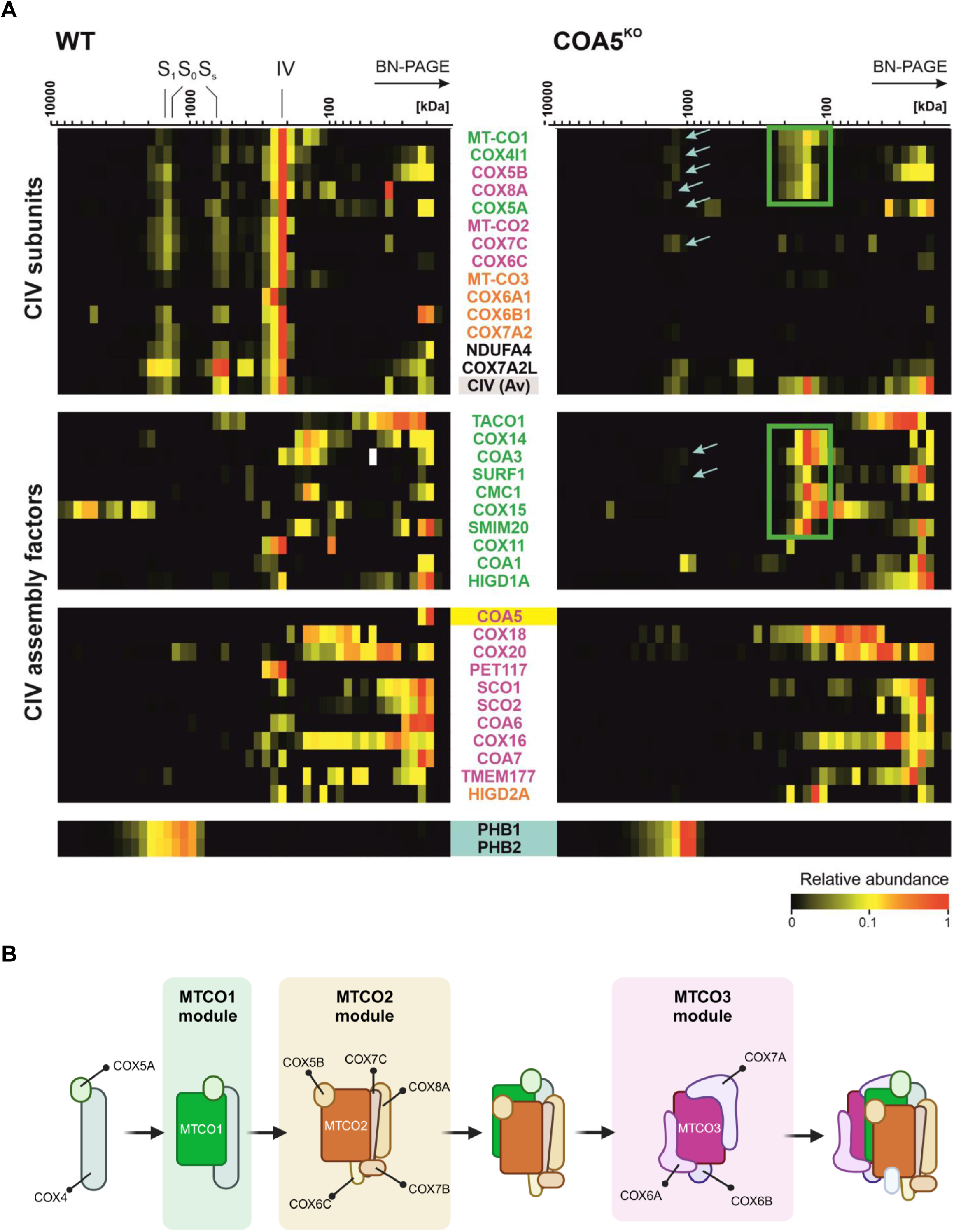
Accumulation of early-stage complex IV assembly intermediate. **(A)** The following heatmap illustrates the distribution of complex IV subunits, assembly factors and the prohibitin complex. The subunits and assembly factors of the assembly modules are cultured as follows: MTCO1 module (green), MTCO2 module (purple), MTCO3 module (orange) and final subunit (black). The light blue arrows indicate subunits that co-migrate with the prohibitin complex. **(B)** A schematic illustration of the sequential assembly intermediates of the CIV holocomplex. Adapted from (Hock *et al*, 2020).

## Discussion

This study identified the previously reported c.157G>C, p.Ala53Pro *COA5* variant in another family. The isolated CIV deficiency associated with the *COA5* variant was observed in the patient fibroblasts and skeletal muscle biopsy, firmly supported by unbiased proteomic profiling of the immortalised *COA5* patient fibroblasts as the biochemical signature of COA5 deficiency. Most importantly, a CRISPR/Cas9 *COA5*^KO^ cell line enabled further interrogation via complexome profiling, narrowing down the involvement of COA5 protein to a specific stage of CIV biogenesis involving MTCO2 stabilisation and its incorporation into the MTCO1-containing subcomplex. This study pinpoints specific questions that arise with regards to the functional role of the COA5 protein as a CIV assembly factor, which we elaborate on further below.

Given that the rare c.157G>C, p.Ala53Pro variant was found only in two incidences where both patients were of identical ethnicity, this could point towards the likelihood of the variant being a founder pathogenic variant within this population. However, this would require further verification through haplotype analysis, necessitating access to patient-derived samples from the previous case. Importantly, overlapping clinical and biochemical phenotypes were observed between the proband in this study and the previously reported patient by Huigsloot and colleagues back in 2019 (Huigsloot *et al*., 2011), further strengthening the claim that the c.157G>C (p.Ala53Pro) *COA5* variant is pathogenic and causative of an isolated mitochondrial complex IV deficiency, associated with loss of steady-state COX proteins and a COX assembly defect in isolation **(Fig 2A-C** and **Fig 3**).

To further characterise the functional impact of COA5, we generated a CRISPR/Cas9-mediated knockout. The *COA5*^KO^ cells successfully replicated the isolated complex IV deficiency observed in the reported patients by indicating a loss of MTCO2 protein but not MTCO1 detected at comparable levels to the isogenic control on SDS-PAGE (**Fig 4A**). While spontaneous degradation of MTCO1 proteins which are unable to be associated with MTCO2 is normally expected, the unaffected steady-state levels of MTCO1 suggest the presence of stabilised MTCO1 subunits despite not being assembled into functional holocomplexes. The loss of the fully assembled complex IV in *COA5*^KO^ cells were not only observed in BN-PAGE analysis but also shown in complexome profiling of the *COA5*^KO^ cells. Most strikingly, this has also been similarly observed in a two-dimensional BN-PAGE of patient fibroblast cell line in the previous report which demonstrated elevated levels of MTCO1 subcomplex but a marked decrease in complex IV holocomplex (Huigsloot *et al*., 2011). Despite this, the remaining OXPHOS complexes (complex I, II, III and V) were unaffected except the unusual accumulation of a protein complex of about 70 kDa detected with SDHA which corresponds to the size of the individual SDHA protein in *COA5*^KO^ cell line.

To further infer the role of COA5 protein and its interacting partner, mitochondrial complexome profiling was employed to dissect co-migrating mitochondrial OXPHOS complexes in the *COA5*^KO^ cell line in comparison to its isogenic control. Notably, the knockout of COA5 protein was confirmed using this method despite the unavailability of a robust antibody for western blotting. Most strikingly, complexome profiling of the *COA5*^KO^ cell line presented evidence of the implication of COA5 in early complex IV assembly, pinpointing its involvement to the stage between MTCO1 maturation and incorporation of MTCO2 into the early assembly intermediate **(Fig 6B)**. This is supported by the following observations: the accumulation of MTCO1-containing subcomplexes (ranging from approximately 100 to 300 kDa) which are absent in the isogenic control, (ii) the apparent absence of MTCO2 subunit and (iii) the loss of fully assembled complex IV (**Fig 6A**). Notably, higher molecular mass complexes were shown containing complex IV subunits in *COA5*^KO^ and could reflect assembly of intermediates with the supercomplex S_0_ scaffold (Fernández-Vizarra & Ugalde, 2022). However, these are more likely to be prohibitin complexes, which direct the free complex IV subunits for degradation rather than supercomplex formation since complex IV biogenesis was completely obstructed (**Fig 6A**) (Back *et al*, 2002; Kohler *et al*, 2023; Steglich *et al*, 1999).

Furthermore, the higher abundance of complex I subunits, which was also observed in the SDS-PAGE analysis of *COA5*^KO^ cell lysates, suggest an accumulation of supercomplex S_0_ (CI+III_2_) which could not form a full respirasome in the absence of CIV holocomplexes. Additionally, the complete loss of supercomplex III_2_+IV reinforced the impact of *COA5*^KO^ resulting in complex IV loss, hence favouring the formation of supercomplexes only containing complexes I and III_2_ to mitigate the OXPHOS defect due to the absence of complex IV. Lastly, the accumulation of free SDHA was not seen in the isogenic control. This may either indicate an upregulation of SDHA as a compensatory mechanism due to the OXPHOS defect or an accumulation of a co-migrating complex of SDHA with CII assembly factors, such as SDHAF1 or SDHAF2 (Martínez-Reyes & Chandel, 2020).

Nevertheless, the question remains as to how the c.157G>C (p.Ala53Pro) *COA5* variant affects mitochondrial CIV assembly. The variant likely affects the CX_9_C motifs of the encoded protein which has been associated with the MIA pathway for IMS localisation despite contradicting theories for its yeast counterpart (Bragoszewski *et al*., 2013; Gladyck *et al*., 2021; Herrmann & Köhl, 2007; Khalimonchuk *et al*., 2008; Longen *et al*., 2009) (**Fig 1B**). The replacement of alanine by proline could potentially introduce a kink to the alpha helices of the protein and consequently hampering correct folding of the protein to facilitate its import (von Heijne, 1991). Consequently, verifying the submitochondrial localisation of COA5 protein would be fundamental in further investigation to inform its functional role associated with an early stage of CIV assembly. Nevertheless, structural studies of the impact of the single amino acid change in *COA5*-encoded protein could also be informative for its potential impact on protein folding and subsequent effect onto its correct localisation within the mitochondria.

If the COA5 protein is localised to the IMS, as of most mitochondrial proteins with twin CX_9_C motifs, it will most likely be involved in the delivery or incorporation of copper atoms into complex IV which takes place in IMS. According to STRING (‘**<u>S</u>**earch **<u>T</u>**ool for **<u>R</u>**etrieval of **<u>In</u>**teracting **<u>G</u>**enes/Proteins’; https://string-db.org/), COA5 protein is predicted to interact with protein partners involved in mitochondrial protein import into IMS and COX copper centre integration, though mostly based on text mining rather than experimental data. Besides, the twin CX_9_C motifs have also been linked to Cu(I) binding activity through the reduction of disulphide bonding between the cysteine residues, especially for characterised protein involved in COX maturation (Horng *et al*, 2004; Horng *et al*, 2005; Stroud *et al*, 2015). Interestingly, Nývltová and colleagues also suggested a role for COA5 in the stabilisation of what the authors termed ‘metallochaperone complexes’, comprising COX-specific copper chaperones and haem biosynthesis enzymes required for the maturation and assembly of COX subunits (Nývltová *et al*, 2022). Although beyond the scope of this manuscript, future experiments should focus on interrogating a direct or indirect role of the COA5 protein in the copper metalation of the MTCO2 subunit which, when affected, leads to destabilisation and eventual degradation of the MTCO2 subunit.

Nevertheless, the possibility for the role of COA5 in the mitochondrial matrix should not be eliminated before its submitochondrial localisation is confirmed. Recently, Peker and colleagues demonstrated a two-step mitochondrial import pathway where substrates of the disulphide relay system, including twin CX_9_C protein family, rely on oxidative folding through the MIA pathway in the IMS prior to their transport into the matrix (Peker *et al*, 2023). Matrix localisation of the protein, however, would point towards an involvement in complex IV biogenesis at the RNA or protein levels. A potential hypothesis is that COA5 could act as a translation factor for MTCO2 expression, similar to the involvement of TACO1 and MITRAC12 (otherwise known as COA3) proteins in the translational regulation of MTCO1 protein synthesis (Mick *et al*, 2012; Richman *et al*, 2016; Weraarpachai *et al*, 2009). Moreover, COA5 could also represent a chaperone or stabilising factor for the MTCO2 subunit to facilitate its incorporation into MTCO1-containing subcomplex and therefore impacting synthesis and stability of the MTCO2 protein specifically.

In conclusion, this study provides insights into a distinct mode of pathological complex IV assembly caused by the assembly factor COA5 which specifically disrupts the transitional stage between MTCO1 maturation and MTCO2 incorporation. We present functional evidence to support a role for human COA5 protein in the early stage of complex IV assembly, corroborating its pathogenicity that can contribute to isolated complex IV deficiency. This warrants further investigation to uncover the structural and functional role of COA5 with new insights into understanding its specific involvement in complex IV biogenesis.

## Materials and Methods

### Whole Exome Sequencing

Trio WES (https://www.exeterlaboratory.com/genetics/genome-sequencing/) was conducted at the Exeter Genomics Laboratory as previously described (Chen *et al*., 2023).

### Histopathological and Biochemical Analyses

10 µm of frozen skeletal muscle sections were used in each assessment. For histopathological studies, H&E staining was employed to determine muscle morphology while sequential COX and SDH histochemistry was used to assess COX activity in muscle fibres. Spectrophotometric measurements of OXPHOS enzyme (complexes I-IV) activities were conducted as described in (Taylor *et al*) relative to citrate synthase activity. Quadruple immunofluorescence assays were carried out by measuring NDUFB8 (CI) and MTCO1 (Reddy *et al*) protein abundance against mitochondrial mass marker, porin using in-house analysis software as outlined in (Taylor *et al*.).

### Cell Culture

Patient and age-matched control fibroblasts, as well as U2OS cells, were cultured in High Glucose Dulbecco’s Modified Eagle Media supplemented with 10% fetal bovine serum, 1X non-essential amino acids, 50 µg/ml penicillin, 50 µg/ml streptomycin and 50 µg/ml uridine.

### SDS-PAGE

Cell pellets were harvested when reaching 80-90% confluency and resuspended in lysis buffer (50 mM Tris-HCl (pH 7.5), 130 mM NaCl, 2 mM MgCl_2_, 1 mM phenylmethanesulphonyl fluoride, 1% (v/v) Nonidet^TM^ P-40 and 1X EDTA free protease inhibitor cocktail). The resuspended cell pellets were incubated on ice for 10 minutes, before harvesting the resultant supernatant from centrifugation at 500 g at 4°C for 5 minutes and the protein concentrations were determined using Bradford assay.

Skeletal muscle homogenates were prepared by grinding 20 mg of frozen muscle section into powder using pestle and mortar in liquid nitrogen and resuspended in radioimmunoprecipitation (RIPA) buffer containing 1% Igepal, 1.5% Triton-X-100, 0.5% sodium deoxycholate, 10mM β-mercaptoethanol, 0.1% SDS, 1 mM PMSF and 1X EDTA-free protease inhibitor cocktail. The resuspensions were subjected to a 45-minute incubation on ice followed by three rounds of 15-second homogenisation. The soluble proteins were extracted by centrifugation at 14 000g at 4°C for 10 minutes and protein concentrations were estimated using Pierce^TM^ BCA Protein Assay Kit.

40 µg of protein extracts were resuspended in 1X Laemmli Sample Buffer and denatured at either 95°C for 5 minutes or 37°C for 15 minutes. The samples were then subjected to 12% SDS-PAGE with the Mini-Protean Tetra Cell system and transferred onto a methanol-activated Immobilon-P Polyvinylidene Fluoride (PVDF) membrane using the Mini Trans-Blot Cell system.

### BN-PAGE

Cells were pelleted and resuspended in cell homogenisation buffer comprising of 0.6 M mannitol, 1 mM EGTA, 10 mM Tris-HCL pH 7.4, 1 mM PMSF and 0.1 % (v/v) bovine serum albumin (BSA). The cell suspensions were subjected to three rounds of 15x homogenisation in Teflon glass homogenisers at 4°C with intermittent differential centrifugation at 400 g for 10 minutes at 4°C to separate cytosolic protein fraction. The mitochondrial fractions were pelleted at 11 000 g for 10 minutes at 4°C and washed in cell homogenisation buffer without BSA.

Approximately 40 mg of skeletal muscle sections were processed and homogenised in Muscle Homogenisation Buffer (250 mM sucrose, 20 mM imidazole hydrochloride and 100 mM PMSF) and homogenised in Teflon glass Dounce homogeniser for 15 to 20 rounds at 4°C. The muscle homogenates were then pelleted at 20 000 g for 10 minutes at 4°C and washed twice with Muscle Homogenisation Buffer before pelleting at 20 000 g for 5 minutes at 4°C.

The final pellets were solubilised in 2% n-dodecyl β-D-maltoside (DDM) and subjected to ultracentrifugation at 100 000 g for 15 minutes at 4°C and the supernatants were extracted for protein concentration determination using Pierce^TM^ BCA Protein Assay Kit. About 10 µg of mitochondrial protein complexes were loaded and separated in the precast Native PAGE™ 4-16% Bis-Tris 1.0 mm Mini Protein Gel in the XCell SureLock Mini-Cell Electrophoresis System based on manufacturer’s protocol. The protein complexes were then immobilised onto an Immobilon-P PVDF membrane using the MiniTrans-Blot Cell system.

### Immunoblotting Analysis

The membranes were blocked in 5% milk for an hour at room temperature before immunoblotting with specific primary antibodies and corresponding HRP-conjugated secondary antibodies as listed: OXPHOS cocktail (ab110411, Abcam), MTCO1 (ab14705, Abcam), MTCO2 (ab110258, Abcam), COXIV (ab14744, Abcam), NDUFB8 (ab110242, Abcam), SDHA (ab14715, Abcam), UQCRC2 (ab14745, Abcam), ATP5A (ab14748, Abcam), GAPDH (600004, ProteinTech), VDAC1 (ab14734, Abcam), Polyclonal Rabbit Anti-Mouse Ig/HRP (P0161, Dako) and Polyclonal Swine Anti-Rabbit Ig/HRP (P0399, Dako).

Finally, resultant signal was detected using SuperSignal™ West Pico PLUS Chemiluminescent Substrate (Thermo Scientific) and analysed with ChemiDoc® XRS+ Imaging Systems and Image Lab Software (BioRad).

### On-beads precipitation and protein digestion

Cell pellets were lysed with 120µl RIPA buffer containing protease and phosphates inhibitors. 10µg of protein from all cell lysates were precipitated with 70% acetonitrile onto magnetic beads (MagReSynAmine, Resyn Biosciences). The proteins were washed on the beads with 100% acetonitrile, 70% ethanol and then resuspended in 50µl 50mM ammoniumbicarbonate containing 10mM DTT for reduction of cysteines. Samples were incubated at 37LC for 40min. Then, to alkylate proteins, 50µl of 30mM IAA in 50mM ammonium bicarbonate was added and samples were incubated at RT in the dark for 30 minutes. 0.5µg trypsin was added to each sample for overnight on-beads protein digestion at 37°C. The resulting peptides were concentrated and desalted on EVOTIPS for mass spectrometry analysis according to the standard protocol from EVOSEP.

### LC-MS/MS analysis

LC-MS/MS analysis was carried out using an EVOSEP one LC system (EVOSEP Biosystems, Denmark) coupled to a timsTOF Pro2 mass spectrometer, using a CaptiveSpray nano electrospray ion source (Bruker Corporation, Germany).

200 ng of digested peptides were loaded onto a capillary C18 column (15 cm length, 150 μm inner diameter, 1.5 μm particle size, EVOSEP, Odense Denmark). Peptides were separated at 40 °C using the standard 30 sample/day method from EVOSEP.

The timsTOF Pro2 mass spectrometer was operated in DIA-PASEF mode. Mass spectra for MS were recorded between m/z 100 and 1700. Ion mobility resolution was set to 0.85–1.30 V·s/cm over a ramp time of 100 ms. The MS/MS mass range was limited to m/z 475-1000 and ion mobility resolution to 0.85-1.27 V s/cm to exclude singly changed ions. The estimated cycle time was 0.95 s with 8 cylces using DIA windows of 25 Da. Collisional energy was ramped from 20eV at 0.60 V s/cm to 59eV at 1.60 V s/cm.

Raw data files from LC-MS/MS analyses were submitted to DIA-NN (version 1.8.1) for protein identification and label-free quantification using the library-free function. The UniProt human database (UniProt consortium, European Bioinformatics Institute, EMBL-EBI, UK) was used to generate library in silico from a human FASTA file. Carbamidomethyl (C) was set as a fixed modification. Trypsin without proline restriction enzyme option was used, with one allowed miscleavage and peptide length range was set to 7-30 amino acids. The mass accuracy was set to 15ppm and precursor false discovery rate (FDR) allowed was 0.01 (1%).

LC-MS/MS data quality evaluation and statistical analysis was done using software Perseus ver 1.6.15.0CRISPR/Cas9 Gene Knockout

Wildtype U2OS cells were resuspended in room-temperature Nucleofector Solution added with Supplement from the Cell Line NucleofectorTM Kit V (Lonza) at a density of 1 × 10^6^ cells per nucleofection reaction. COA5-targeting sgRNA (Sigma) was incubated at room temperature with HiFi Cas9 nuclease at a 1:1.2 molar ratio to form ribonucleoprotein complexes. The sgRNA sequence designed to mediate CRISPR/Cas9 knockout of the COA5 gene is as follows: 5’-TTTTGAGTGTAAAAGATCAG-3’. The sgRNA-Cas9 RNP complexes were nucleofected into wildtype U2OS cells on the NucleofectorTM 2b Device (Lonza). Nucleofected cells were resuspended in growth media as outlined in Section 2.6.1 before transferring to a 6-well plate. The cells were incubated at 37°C, 5% CO2 for 48 hours before isolating into single cell clones using FACSAria™ Fusion Flow Cytometer (BD Biosciences) in four 96-well plates. Sanger sequencing chromatographs of the selected single cell clones were analysed using Inference of CRISPR Edit (ICE) analysis (Synthego; https://ice.synthego.com/) to identify isogenic controls and COA5 knockout cell line.

### Complexome Profiling (BN-PAGE, Trypsin Digestion, LC-MS/MS, Quantification)

Enriched mitochondrial proteins were extracted from COA5^KO^ and isogenic control cell lines and solubilised with digitonin as described in (Giese et al., 2021). Equal amount of the solubilised mitochondrial protein extracts were subjected to a 3 to 18% acrylamide gradient gel (14cm ×14cm) for BN-PAGE as outlined in (Wittig, Braun and Schägger, 2006). The gel was then stained with Coomassie blue and cut into equal fractions then transferred to 96-well filter plates. The gel fractions were then destained in 50 mM ammoniumbicarbonate (ABC) followed by protein reduction using 10 mM DTT and alkylation in 20 mM iodoacetamide. Protein digestion was carried out in digestion solution (5 ng trypsin/μl in 50 mM ABC, 10% acetonitrile (ACN), 0.01% (w/v) ProteaseMAX surfactant (Promega), 1 mM CaCl_2_) at 37°C for at least 12 hours. After the recovery in the new 96-well plate, the peptides were dried in a SpeedVac (ThermoFisher) and finally resuspended in 1% ACN and 0.5% formic acid. Nano liquid chromatography and mass spectrometry (nanoLC/MS) was carried out on Thermo Scientific™ Q Exactive Plus equipped with ultrahigh performance liquid chromatography unit Dionex Ultimate 3000 (ThermoFisher) and a Nanospray Flex Ion-Source (ThermoFisher). The MS data was analysed using MaxQuant software at default settings and the recorded intensity-based absolute quantifications (iBAQ) values were normalised to isogenic control cell line.

## Data Availability

The mass spectrometry proteomics data for label-free whole cell proteomics and complexome profiling produced in this study have been deposited to the ProteomeXchange Consortium via the PRIDE partner repository (Perez-Riverol *et al*, 2022) and assigned the dataset identifier PXD050891 and PXD053461 respectively.

## Acknowledgements

RWT is supported by the Wellcome Centre for Mitochondrial Research (203105/Z/16/Z), the Medical Research Council (MRC) International Centre for Genomic Medicine in Neuromuscular Diseases (MR/S005021/1), the UK NIHR Biomedical Research Centre in Age and Age-Related Diseases award to the Newcastle upon Tyne Hospitals NHS Foundation, the Lily Foundation, LifeArc and the UK NHS Highly Specialised Service for Rare Mitochondrial Disorders. RWT and MW are supported by the Pathology Society; RWT, MW and AP are supported by Mito Foundation. JXT was supported by a PhD studentship from the Lily Foundation and a Newcastle University Overseas Research Studentship award. CBJ is supported by funding from the Academy of Finland (decision #336455), the Magnus Ehrnroot Foundation and the Jane and Aatos Erkko Foundation (#230004). The Proteomics Core Facility, University of Oslo/Oslo University Hospital is supported by the Core Facilities program of the South-Eastern Norway Regional Health Authority, and is a member of the National Network of Advanced Proteomics Infrastructure (NAPI) which is funded by the Research Council of Norway INFRASTRUKTUR-program (project number: 295910). IW is supported by the Deutsche Forschungsgemeinschaft (DFG): SFB1531-S01, project number 456687919 and WI 3728/3-1, project number 515944830.

## Author Contributions

**Conceptualization**: AP, MW, IW and RWT

**Data curation and formal analysis:** JXT, ACO, JM, LST, GM, SKS, AS, KS, LH, SH, TAN

**Clinical care of the family:** JD

**Supervision:** AP, MW, CBJ, IW and RWT

**Drafting the manuscript:** JXT, AP, MW and RWT

**Critical revision and editing of the manuscript:** all authors

**Funding acquisition:** AP, MW, IW and RWT

**Disclosure and competing interests statement:** The authors declare no conflicts of interest.

## The Paper Explained

### PROBLEM

A homozygous *COA5* variant was previously reported to cause a mitochondrial cardiomyopathy phenotype but the role of the COA5 protein is poorly defined despite being proposed to be an assembly factor for cytochrome *c* oxidase (COX), an essential protein of the mitochondrial respiratory chain.

### RESULTS

Through a series of biochemical experiments including LC-MS/MS-based complexome profiling, our study showed that the human COA5 protein is essential for the biogenesis and incorporation of the MTCO2 subunit, specifically, to form functional COX holocomplexes.

Additionally, we present an additional case of COA5-related mitochondrial disease, confirming the pathogenicity of the reported homozygous *COA5* variant that leads to mitochondrial dysfunction.

### IMPACT

We have provided additional insights into the functional role of human COA5 protein and its specific implications on early-stage COX assembly, correlating to isolated COX deficiencies observed in patients harbouring a pathogenic, homozygous *COA5* variant.

---

**Figure EV1.**
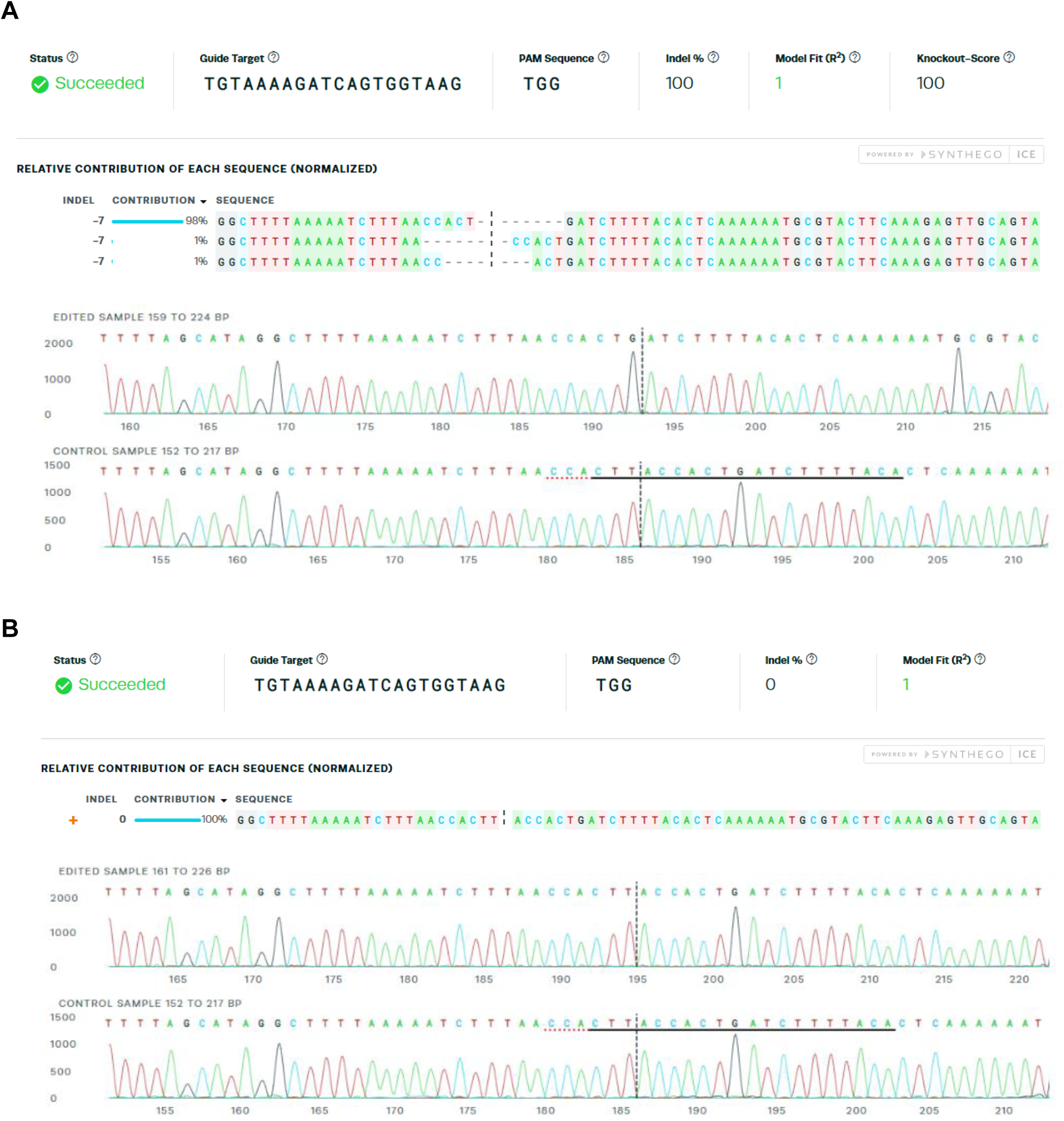
Synthego Inference of CRISPR Edits (ICE) analysis of *COA5* CRISPR knockout and isogenic control cell lines. **(A)** A homozygous 7bp deletion (c.287_290+3del, p.Val61del) around the expected cut site of the expected cut site of the *COA5*-targeting sgRNA (indicated with black dotted line) was verified, generating a full knockout of the *COA5* gene. Sanger sequencing traces in the bottom panel illustrate the deletion of seven bases in the *COA5*^KO^ cells compared to wildtype sequence shown below which contains protospacer adjacent motif (PAM) site underlined in red dotted line and the sgRNA binding site in black. **(B)** Wildtype sequence was confirmed through Sanger sequencing in the isogenic control cell line selected from the cell population that was nucleofected with Cas9 and sgRNA targeting the *COA5* gene.

**Figure EV2.**
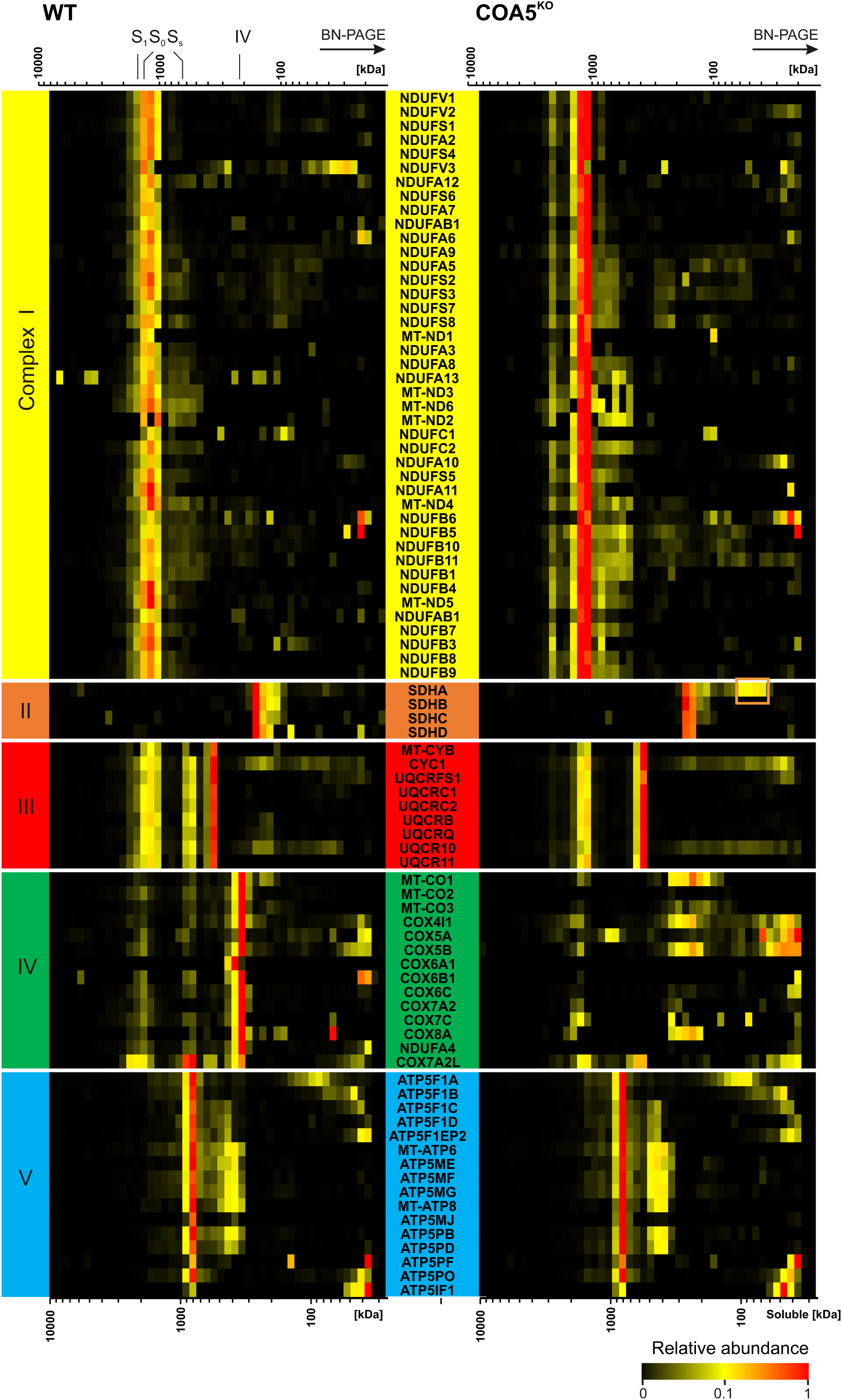
Mitochondrial complexome profiling of *COA5*^KO^ and wildtype (COA5 WT) isogenic control cell lines. The data are presented as a heat map, corresponding to protein components of individual OXPHOS complexes. The size of the complexes ranges from 10000 kDa to 10 kDa (from left to right). The corresponding complexes were highlighted above the heatmap as individual subunits, fully assembled complex IV holocomplexes (IV), complex III dimers (III_2_), small supercomplex of complex III dimers and complex IV (S_s_, III_2_+IV) and supercomplexes containing complex I, complex III dimers (S_0_) and complex IV (S_1_). Relative abundance of each protein was represented from low to high according to colour scale illustrated on the bottom right.

**Figure EV3.**
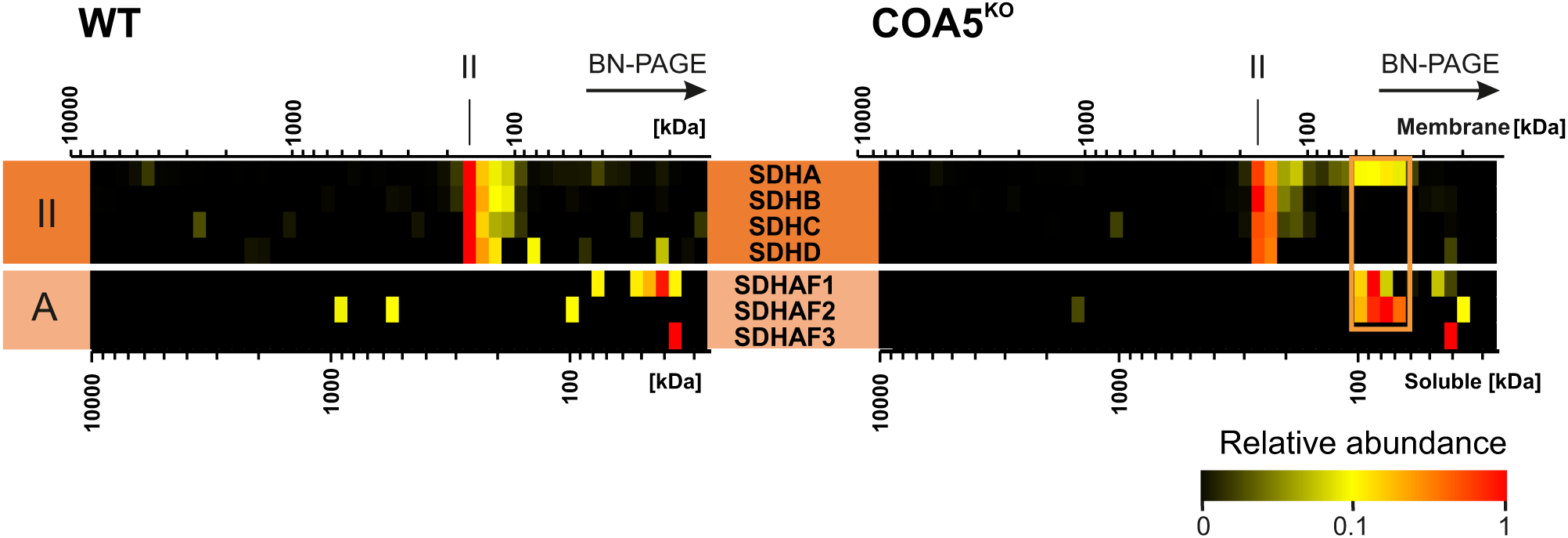
Complex II assembly intermediates in COA5 KO. The relative protein abundance is presented as a heat map, corresponding to protein components of complex II. The size of the complexes ranges from 10000 kDa to 10 kDa (from left to right) and are given as native membrane (top scale) and soluble (lower scale) protein calibration. The orange box indicates an assembly intermediate in the COA5^KO^ cell line.

